# Assessing the Feasibility of Processing a Paper-based Multilingual Social Needs Screening Questionnaire Using Artificial Intelligence

**DOI:** 10.1101/2024.03.25.24304755

**Authors:** Obinna I. Ekekezie

**Affiliations:** Cambridge Health Alliance, Cambridge, MA, United States of America; Harvard Medical School, Boston, MA, United States of America

## Abstract

The collection of Social Determinants of Health (SDoH) data is increasingly mandated by healthcare payers, yet traditional paper-based methods pose challenges in terms of cost effectiveness, accuracy, and completeness when manually entered into Electronic Health Records (EHRs). This study explores the application of artificial intelligence (AI), specifically using a document understanding model (Microsoft Azure Document Intelligence) and large language models (OpenAI’s GPT-4 Turbo and GPT-3.5 Turbo), to automate the conversion of paper-based Social Determinants of Health (SDoH) questionnaires into structured, machine-readable formats that could theoretically be incorporated into EHRs. Using a dataset of synthetic and scanned examples, the study compares the performance of the GPT-3.5 and 4 Turbo base models and fine-tuned GPT-3.5 Turbo models on this task. Findings indicate that GPT-4 Turbo outperforms GPT-3.5 Turbo in accuracy and consistency, with fine-tuning enhancing GPT-3.5 Turbo’s performance and consistency in several languages. These results suggest that AI could prove to be an accurate alternative to manual data entry, with important implications for improving how SDoH data is incorporated into EHRs. Future research should address data privacy, security concerns, cost considerations, and the technical aspects of incorporating AI-generated data into EHRs.

**Description:** This study explores the application of artificial intelligence (AI), specifically using a document understanding model (Microsoft Azure Document Intelligence) and large language models (OpenAI’s GPT-4 Turbo and GPT-3.5 Turbo), to automate the conversion of paper-based Social Determinants of Health (SDoH) questionnaires into structured, machine-readable formats that could theoretically be incorporated into a patient’s electronic health records (EHRs).

## Introduction

In recent years, healthcare payers have been incentivizing or even mandating the collection of information about the social determinants of health (SDoH). These refer to the social and economic factors, such as social isolation, interpersonal violence, or lack of access to housing, transportation, or food, that can impact the health and well-being of individuals^1,2^.

Like many other forms of personal or demographic information asked of patients, this data is often collected using paper-based questionnaires that are later manually entered into an electronic health record (EHR) by clinicians or non-clinicians^2^. In addition to manual data entry being expensive, it is worth noting that previous studies have found that manually entered data is often less accurate and less complete than information that is extracted using automations^3^. Some of these issues can be mitigated by administering the questionnaires in an electronic format using tablets and other devices. However, developing such capabilities can require deep technical expertise as well as substantial investments of time and resources that many healthcare providers may not have^4^.

With recent advancements in artificial intelligence (AI), particularly with respect to document understanding models and using large language models (LLMs) for natural language processing (NLP) tasks, it is now more feasible than ever to extract structured data from documents^5,6^. Being able to reliably and efficiently extract structured data from paper-based questionnaires, could help more healthcare providers meet these SDoH-related mandates. Previous research has explored using transformer-based, deep learning NLP methods to analyze questionnaire responses; however, this research did not employ document understanding models or LLMs and assumed the questionnaire was already in a structured format^7^. This study sought to explore the feasibility of using a document understanding model in concert with an LLM to generate a structured representation of a paper-based SDoH questionnaire response.

## Methods

### Document Understanding Model

A document understanding model uses AI to comprehend, classify, and extract meaningful information from unstructured or semi-structured documents^5^. For this study, both Microsoft Azure Document Intelligence and Google Document AI were considered as potential document understanding models. Ultimately, Document Intelligence was chosen due to Document AI being less reliable in detecting checkboxes in documents as per the Google Document AI July 18, 2023 release notes.

The Document Intelligence Layout Model application programming interface (API) was used to process the scanned questionnaires as both text and markdown, a markup language used to format plain text. The text and markdown included special selection mark tokens denoting whether a checkbox was “selected” or “unselected” (Fig. S1).

### Large Language Model (LLM)

OpenAI’s GPT-4 Turbo (“gpt-4-turbo-2024-04-09”), which is amongst the best LLMs currently available^8^, was used as a benchmark. As it is currently significantly less expensive than GPT-4 Turbo, GPT-3.5 Turbo (“gpt-3.5-turbo-0125”) was chosen for conducting the experiments. It was also chosen for conducting experiments since OpenAI provides a service that allows developers to fine-tune the base model on specific tasks. Fine-tuning involves making small adjustments to a pre-trained model’s parameters, optimizing it for specific types of data or tasks^9^. It is worth noting that training a fine-tuned version of an OpenAI model has costs and that fine-tuned models are more expensive to use than base models. Finally, in order to make the LLMs’ outputs more deterministic, the temperature was set to zero and a seed was used. JavaScript Object Notation (JSON) mode was also utilized to ensure the LLMs’ responses were formatted as valid JSON.

### Dataset

The SDoH questionnaire used in this feasibility study was created by Cambridge Health Alliance (CHA) and is based on existing research^10–13^. To meet the needs of its diverse patient population, the CHA SDoH questionnaire is available in eight languages: Arabic, Chinese, English, Haitian Creole, Hindi, Nepali, Portuguese, and Spanish. Regardless of the language, each questionnaire contains 10 questions and a total of 26 different answer choices each represented by a checkbox. To avoid exposing any personal health information (PHI), a synthetic dataset was created (Fig. 1). So that it would be possible to explore how well the models generalized to different inputs, a second dataset of test examples was created by filling out questionnaires by hand, scanning them as PDFs, and then manually labeling them (for more detailed information, see “Creating the Datasets” and “Analyzing the Text and Markdown Extracted from the Synthetic and Scanned Examples” in the Supplementary Appendix).

**Figure 1.**
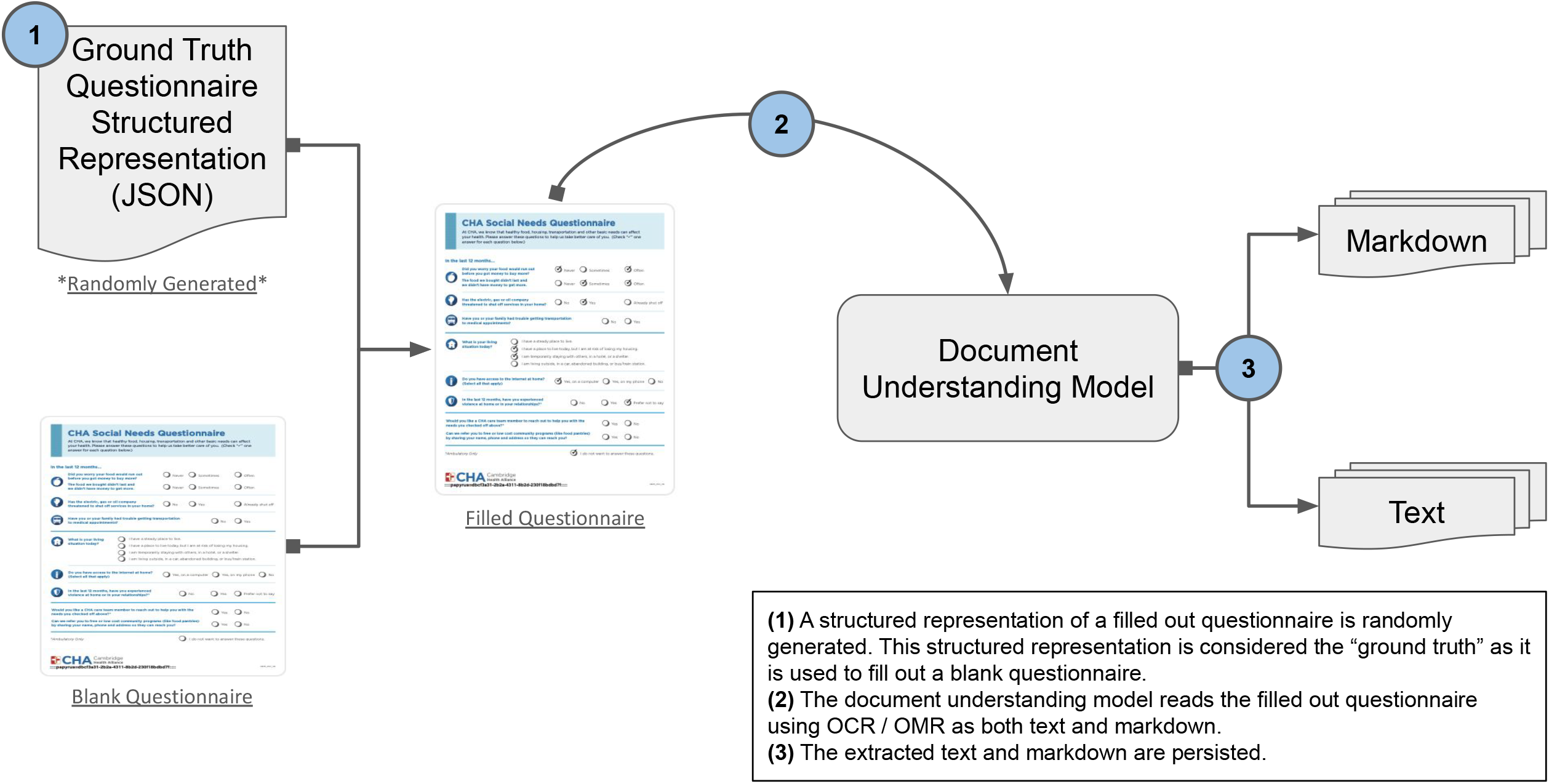
Creating Synthetic Examples from a Ground Truth

### Experiments

The LLM’s task was to create a structured JSON representation of a scanned, filled out questionnaire. DSPy^14^, a Python programming framework designed for programming with LLMs, was used to run the experiments (Fig. 2). Two different DSPy programs were tested: one that utilized chain of thought, a method where the model is prompted to articulate intermediate steps or reasoning before providing a final answer^15^, and another that utilized a zero-shot approach, where the model attempts to perform the task without any prior examples specific to that task^9^. In addition, three combinations of program inputs were tested: text only, markdown only, and both text and markdown. To increase the likelihood of the LLM producing an output that adhered to the structured representation’s schema, the LLM’s program inputs also included a structured representation of a blank questionnaire it was instructed to use as a template.

**Figure 2.**
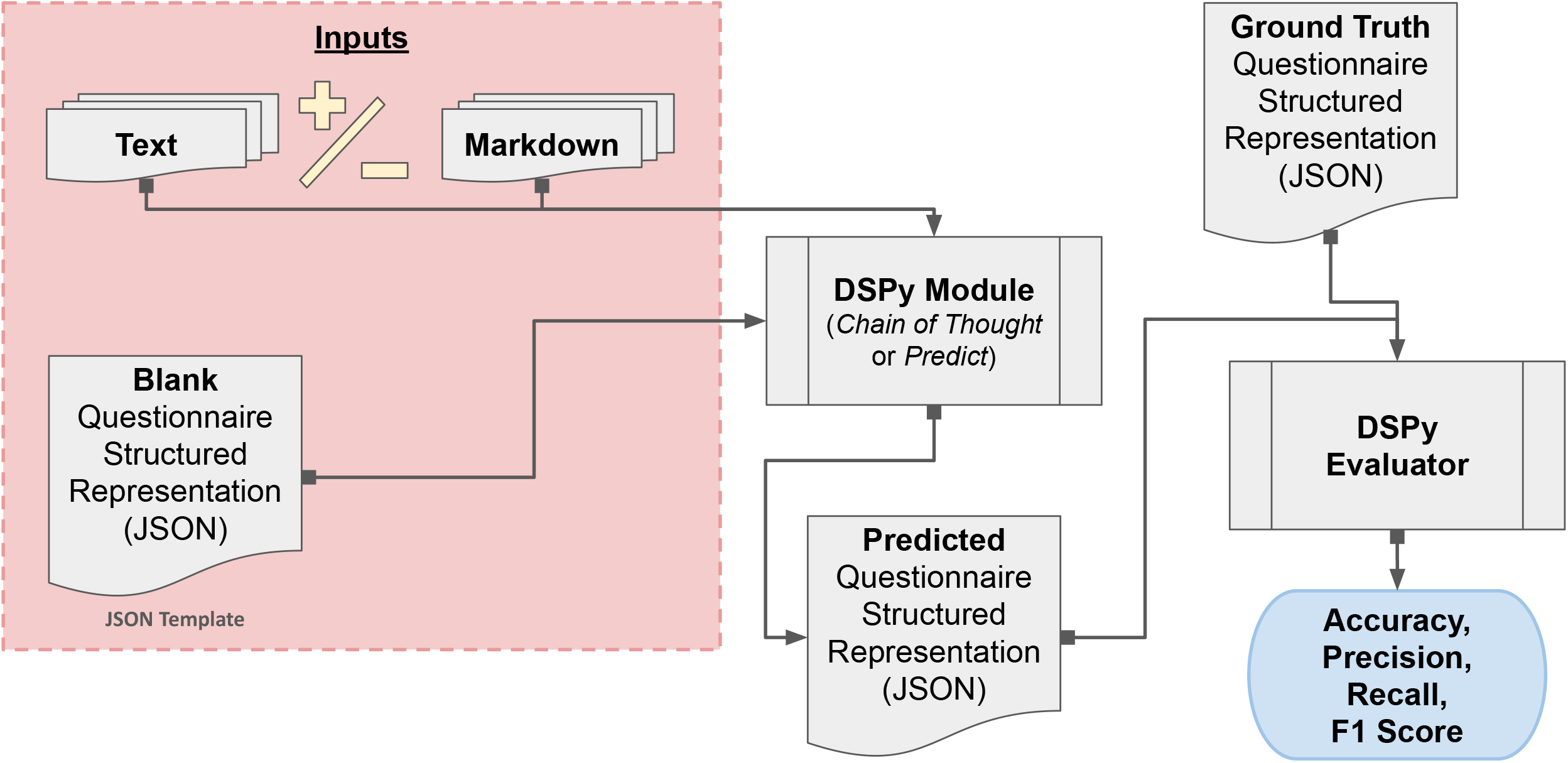
Configuring DSPy Programs and Running Experiments

Each program’s performance was measured by comparing the LLM’s output (i.e. prediction) against a ground truth and calculating the accuracy, precision, recall, and F1 score. It is important to note that all four metrics were zero if the prediction was invalid JSON or failed to adhere to the structured representation’s expected schema. Lastly, all experiments were run three times to assess the LLMs’ consistency.

### Fine-tuning GPT-3.5 Turbo

Given that OpenAI generally recommends using 50 to 100 fine-tuning examples as well as to use a separate set of examples for validation, the fine-tuning dataset was created by identifying the 16 most challenging training examples with the lowest F1 scores for each language (i.e., a total of 128 examples in the smaller fine-tuning dataset). 13 of these challenging examples per language were reserved for fine-tuning, while the remaining three were reserved for validating the fine-tuned model. To understand whether using more examples for fine-tuning might improve performance, the fine-tuning process was repeated using the 32 most challenging training examples for each language (i.e., a total of 256 examples in the larger fine-tuning dataset), reserving 26 examples for training the fine-tuned model and the remaining six for validation. The fine-tuned GPT-3.5 Turbo model with 128 examples is referred to as “small” while the one that was trained on 256 examples is referred to as “large” (see “Fine-Tuning GPT-3.5 Turbo” and Table S1 in the Supplementary Appendix for additional information).

## Results

### Program Module and Inputs Combinations

Based on the results of comparing the different program modules and inputs combinations on the synthetic training dataset, the program using a zero shot approach with both markdown and text inputs yielded the best results (Fig. 3). Consequently, it was chosen for additional fine-tuning using both the smaller and then larger subsets of 128 and 256 challenging training examples, respectively, as described above.

**Figure 3.**
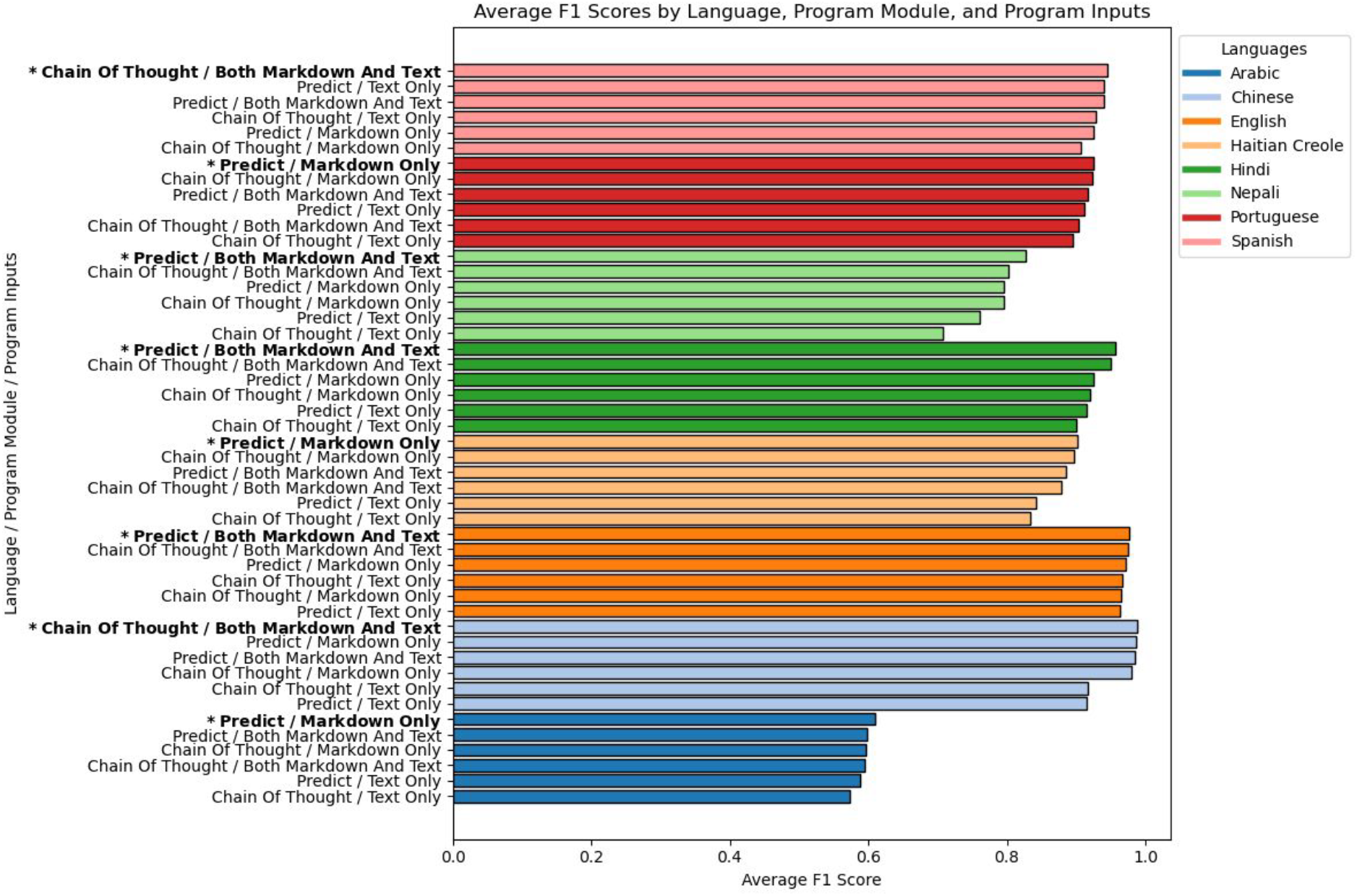
Identifying Best Performing Program Module / Inputs Combination

### Synthetic Examples Test Dataset

Comparing the performance of the four models on the synthetic test dataset (Table 1 and Fig. 4) showed that GPT-4 Turbo’s baseline performance was superior to GPT-3.5 Turbo’s as it substantially outperformed the latter with respect to accuracy, precision, recall, and F1 score across each language (though there was an exception in which the precision for English test examples was 0.991 and 0.988 for GPT-3.5 and 4 Turbo, respectively). The fine-tuned models generally outperformed the GPT-3.5 and 4 Turbo base models. The small fine-tuned model seemed to perform better on Arabic and Nepali test examples than the large fine-tuned model did. In addition, overall, it appears as though using more examples for fine-tuning seemed to degrade performance slightly for most languages. That being said, in the case of Portuguese examples, it appeared as though the performance improved when using the larger subset of challenging training examples for fine-tuning. As illustrated by the lower amount of variation, it appears as though the fine-tuned models produced the same output when given the same input more often than the base models did (see “Comparing Performance on Synthetic Testing Dataset” in the Supplementary Appendix for additional information).

**Table 1:** Performance by Model by Language on Synthetic and Scanned Test Examples *Note: Metrics are mean accuracy, mean precision, mean recall, and mean F1 score*.

| <u>Model Name and Testing Dataset</u> | <u>Language</u> | <u>Accuracy</u> | <u>Precision</u> | <u>Recall</u> | <u>F1 Score</u> |
| --- | --- | --- | --- | --- | --- |
| GPT-3.5 Turbo (Base Model)<br>**Synthetic Examples Dataset** | Arabic | 0.594 | 0.590 | 0.462 | 0.513 |
|  | Chinese | 0.979 | 0.978 | 0.975 | 0.976 |
|  | English | 0.978 | 0.991 | 0.973 | 0.981 |
|  | Haitian Creole | 0.896 | 0.887 | 0.886 | 0.886 |
|  | Hindi | 0.937 | 0.901 | 0.942 | 0.920 |
|  | Nepali | 0.808 | 0.827 | 0.740 | 0.764 |
|  | Portuguese | 0.915 | 0.913 | 0.914 | 0.913 |
|  | Spanish | 0.947 | 0.951 | 0.930 | 0.939 |
|  | **Overall** | **0.882** | **0.880** | **0.853** | **0.862** |
| GPT-3.5 Turbo Fine-Tuned (Small)<br>**Synthetic Examples Dataset** | Arabic | 0.795 | 0.854 | 0.688 | 0.759 |
|  | Chinese | 1.000 | 1.000 | 1.000 | 1.000 |
|  | English | 0.985 | 0.988 | 0.988 | 0.988 |
|  | Haitian Creole | 0.996 | 1.000 | 0.988 | 0.993 |
|  | Hindi | 1.000 | 1.000 | 1.000 | 1.000 |
|  | Nepali | 0.973 | 0.976 | 0.965 | 0.970 |
|  | Portuguese | 0.931 | 0.929 | 0.929 | 0.929 |
|  | Spanish | 1.000 | 1.000 | 1.000 | 1.000 |
|  | **Overall** | **0.960** | **0.968** | **0.945** | **0.955** |
| GPT-3.5 Turbo Fine-Tuned (Large)<br>**Synthetic Examples Dataset** | Arabic | 0.774 | 0.816 | 0.660 | 0.727 |
|  | Chinese | 1.000 | 1.000 | 1.000 | 1.000 |
|  | English | 0.985 | 0.994 | 0.976 | 0.984 |
|  | Haitian Creole | 0.992 | 1.000 | 0.979 | 0.989 |
|  | Hindi | 0.987 | 1.000 | 0.973 | 0.985 |
|  | Nepali | 0.940 | 0.967 | 0.891 | 0.923 |
|  | Portuguese | 0.962 | 0.958 | 0.958 | 0.958 |
|  | Spanish | 0.982 | 0.992 | 0.972 | 0.981 |
|  | **Overall** | **0.953** | **0.966** | **0.926** | **0.943** |
| GPT-4 Turbo (Base Model)<br>**Synthetic Examples Dataset** | Arabic | 0.681 | 0.693 | 0.619 | 0.650 |
|  | Chinese | 1.000 | 1.000 | 1.000 | 1.000 |
|  | English | 0.983 | 0.985 | 0.985 | 0.985 |
|  | Haitian Creole | 0.983 | 0.989 | 0.982 | 0.986 |
|  | Hindi | 0.994 | 0.998 | 0.978 | 0.987 |
|  | Nepali | 0.896 | 0.875 | 0.879 | 0.876 |
|  | Portuguese | 1.000 | 1.000 | 1.000 | 1.000 |
|  | Spanish | 0.978 | 0.979 | 0.975 | 0.976 |
|  | **Overall** | **0.939** | **0.940** | **0.927** | **0.932** |
| GPT-3.5 Turbo (Base Model)<br>**Scanned Examples Dataset** | Arabic | 0.637 | 0.546 | 0.411 | 0.464 |
|  | Chinese | 1.000 | 1.000 | 1.000 | 1.000 |
|  | English | 0.991 | 1.000 | 0.978 | 0.988 |
|  | Haitian Creole | 0.816 | 0.725 | 0.765 | 0.744 |
|  | Hindi | 0.885 | 0.853 | 0.844 | 0.849 |
|  | Nepali | 0.876 | 0.884 | 0.765 | 0.813 |
|  | Portuguese | 0.889 | 0.836 | 0.870 | 0.852 |
|  | Spanish | 0.940 | 0.926 | 0.904 | 0.914 |
|  | **Overall** | **0.879** | **0.846** | **0.817** | **0.828** |
| GPT-3.5 Turbo Fine-Tuned (Small)<br>**Scanned Examples Dataset** | Arabic | 0.372 | 0.250 | 0.207 | 0.227 |
|  | Chinese | 0.974 | 1.000 | 0.930 | 0.963 |
|  | English | 0.987 | 1.000 | 0.963 | 0.980 |
|  | Haitian Creole | 1.000 | 1.000 | 1.000 | 1.000 |
|  | Hindi | 0.987 | 1.000 | 0.967 | 0.982 |
|  | Nepali | 0.949 | 0.963 | 0.893 | 0.926 |
|  | Portuguese | 0.936 | 0.900 | 0.933 | 0.916 |
|  | Spanish | 1.000 | 1.000 | 1.000 | 1.000 |
|  | **Overall** | **0.901** | **0.889** | **0.862** | **0.874** |
| GPT-3.5 Turbo Fine-Tuned (Large)<br>**Scanned Examples Dataset** | Arabic | 0.697 | 0.708 | 0.389 | 0.485 |
|  | Chinese | 0.987 | 1.000 | 0.963 | 0.980 |
|  | English | 1.000 | 1.000 | 1.000 | 1.000 |
|  | Haitian Creole | 1.000 | 1.000 | 1.000 | 1.000 |
|  | Hindi | 0.987 | 1.000 | 0.967 | 0.982 |
|  | Nepali | 0.889 | 0.960 | 0.726 | 0.825 |
|  | Portuguese | 0.949 | 0.930 | 0.933 | 0.930 |
|  | Spanish | 1.000 | 1.000 | 1.000 | 1.000 |
|  | **Overall** | **0.939** | **0.950** | **0.872** | **0.900** |
| GPT-4 Turbo (Base Model)<br>**Scanned Examples Dataset** | Arabic | 0.628 | 0.503 | 0.519 | 0.509 |
|  | Chinese | 1.000 | 1.000 | 1.000 | 1.000 |
|  | English | 1.000 | 1.000 | 1.000 | 1.000 |
|  | Haitian Creole | 0.936 | 0.893 | 0.926 | 0.908 |
|  | Hindi | 0.974 | 0.970 | 0.967 | 0.967 |
|  | Nepali | 0.786 | 0.702 | 0.702 | 0.702 |
|  | Portuguese | 0.987 | 0.967 | 1.000 | 0.982 |
|  | Spanish | 0.970 | 0.975 | 0.942 | 0.958 |
|  | **Overall** | **0.910** | **0.876** | **0.882** | **0.878** |

**Figure 4.**
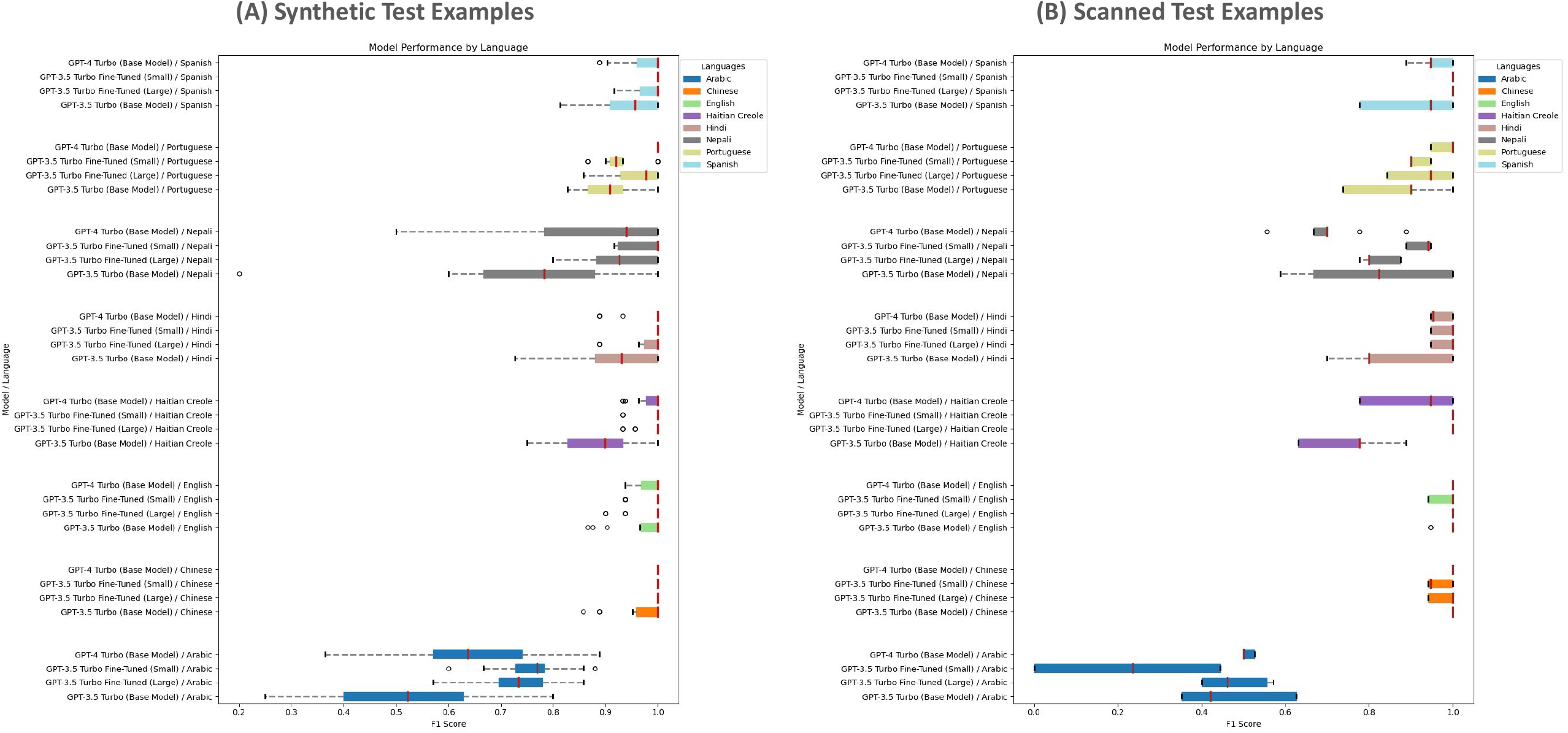
Performance (F1 Score) by Language and Model

### Scanned Examples Test Dataset

Testing the model on the test dataset derived from scanning questionnaires that had been manually filled out by hand yielded different results (Table 1 and Fig. 4). In this case, the large fine-tuned model had the best overall performance suggesting that including more fine-tuning examples may have helped the model generalize better. Of note, the small fine-tuned model’s performance on Arabic, Chinese, and English examples degraded relative to the other models with the most significant degradation in performance occurring with Arabic examples. It was the only model that failed to always produce predictions that adhered to the questionnaire’s structured representation’s schema. As was observed when testing using synthetic examples, the fine-tuned models predictions generally varied less than the base models’ (see “Comparing Performance on Scanned Testing Dataset” in the Supplementary Appendix for additional information).

## Discussion

There were a number of languages, including Chinese, English, Haitian Creole, Hindi, Portuguese, and Spanish, for which at least one of the models tested achieved both an accuracy and F1 score of at least 0.95. It is also worth noting that, without fine-tuning, the GPT-3.5 Turbo base model was able to achieve this level of performance only for English and Chinese examples initially. Moreover, despite additional fine-tuning, GPT-3.5 Turbo still performed poorly on Arabic and Nepali examples, and it somewhat struggled with Portuguese examples but less so. These results suggest AI could soon be used to transform paper-based questionnaires into structured, machine-readable formats for inclusion in EHRs for many of the most commonly spoken languages in the United States^16^. However, it is important to note that the degree to which and direction in which fine-tuning impacted performance across languages was not uniform and that improved performance in one language sometimes coincided with worsening performance in another. This observation highlights the importance of testing any AI model after undergoing additional training to be sure the model’s performance actually improved and that it did not regress in other important ways^17^.

Previous research indicates that administering these questionnaires electronically using tablets in primary care clinics can improve screening quality, and that requiring manual entry of paper-based questionnaire responses into EHRs can potentially compromise screening effectiveness^18^. Employing AI to automate data extraction from paper-based questionnaires could enhance the efficiency and quality of SDoH screening in clinics lacking the necessary devices and technical expertise and reduce the disparity between the clinics that do and do not have these capabilities as well.

Although using AI for generating structured representations of paper-based questionnaires is promising, there could be some concerns with sharing sensitive information like a patient’s social needs with the third parties that own these AI models. Additionally, it is crucial to evaluate potential security threats healthcare organizations employing LLMs could face, such as the risk of prompt injection attacks where attackers hijack LLMs with malicious instructions^19^. Furthermore, integrating the LLM-generated structured representation into the EHR would necessitate post-processing to convert it into an EHR-compatible format, e.g. the HL7 Fast Healthcare Interoperability Resources (FHIR) standard^20^, and additional validation to ensure the output’s integrity. It would also be worthwhile to explore the degree to which the cost differential between using manual data entry versus using AI could be impacted by running such an application on HIPAA-compliant infrastructure as would be required if real patient data were used. That said, the results thus far are quite promising and suggest that it would be worthwhile to address these considerations in future research.

## Data Availability

All data produced in the present study are available upon reasonable request to the author.
In addition, a GitHub repo containing the results from assessing GPT-3.5 Turbo on the training synthetic examples and from comparing models on the two testing datasets (synthetic and scanned) as well as the LLM calls and responses is available. The repo also contains the notebooks (HTML) referenced in the Supplementary Appendix.

https://github.com/oekekezie/poc-ai-sdoh-questionnaire-v2/

## Acknowledgements

Lara Jirmanus, MD, MPH and the rest of the team involved in spearheading the efforts to implement screening for social needs in primary care at Cambridge Health Alliance. Ikechukwu Ekekezie, MD, Hannah Galvin, MD, FAAP, FAMIA, CHCIO, Hsiang Huang, MD, MPH, Thomas Seufert, MD, and Rajendra Aldis, MD, MS for providing constructive feedback regarding the final draft of this article.

