## Supplementary Appendix for "Assessing the Feasibility of Processing a Paper-based Multilingual Social Needs Screening Questionnaire Using Artificial Intelligence"

Obinna I. Ekekezie, M.D.[a,b]

[a] *Cambridge Health Alliance, Cambridge, MA, United States of America*
[b] *Harvard Medical School, Boston, MA, United States of America*

### Figure S1: Text Extracted by Document Intelligence Layout API

The text extracted from a questionnaire by the Document Intelligence Layout API includes special selection mark tokens, “:selected:” or “:unselected:”, denoting whether a checkbox was checked or unchecked, respectively:

*CHA Social Needs Questionnaire*
*At CHA, we know that healthy food, housing, transportation and other basic needs can affect your health.*
*Please answer these questions to help us take better care of you. (Check "V" one answer for each question below.)*
*In the last 12 months ...*
*Did you worry your food would run out before you got money to buy more? :selected: Never :unselected:*
*Sometimes :unselected: Often*
*The food we bought didn't last and we didn't have money to get more. :selected: Never :selected:*
*Sometimes :selected:*
*Often*
*Has the electric, gas or oil company threatened to shut off services in your home? :unselected: No :selected:*
*Yes :unselected: Already shut off*
*Have you or your family had trouble getting transportation to medical appointments? :unselected: No*
*:selected:*
*Yes*
*What is your living situation today? :unselected:*
*m I have a steady place to live. m I have a place to live today, but I am at risk of losing my housing. m*
*:unselected: :selected: I am temporarily staying with others, in a hotel, or a shelter. :unselected:*
*m I am living outside, in a car, abandoned building, or bus/train station.*
*i Do you have access to the internet at home? (Select all that apply) :selected: Yes, on a computer*
*:unselected: Yes, on my phone :selected: No*
*In the last 12 months, have you experienced violence at home or in your relationships ?* :unselected: No*
*:selected:*
*Yes :selected:*
*Prefer not to say*
*Would you like a CHA care team member to reach out to help you with the needs you checked off above ?**
*:selected: Yes :unselected: No*
*Can we refer you to free or low cost community programs (like food pantries) by sharing your name, phone and address so they can reach you? :unselected: Yes :unselected: No*
**Ambulatory Only :unselected: I do not want to answer these questions.*
*CHA*
*Cambridge Health Alliance*
*:::: papyrus=dbcf3a31-2b2a-4311-8b2d-230f18bdbd7f ::::*
*GR20_200_SN :selected:*

#### Creating the Datasets

To avoid exposing any personal health information (PHI), a synthetic dataset was created. For each of the eight languages (Arabic, Chinese, English, Haitian Creole, Hindi, Nepali, Portuguese, and Spanish), 50 synthetic examples were generated by randomly generating structured representations of completed questionnaires (Fig. S2) that were then used to render PDFs of the questionnaire. The PDFs were then processed using the Document Intelligence Layout API and the extracted text and markdown were persisted (see "Analyzing the Text and Markdown Extracted from the Synthetic and Scanned Examples" in the Supplementary Appendix for additional information).

For each language, 40 examples were allocated to the training dataset which was used to assess GPT-3.5 Turbo's baseline performance on the task. The remaining 10 examples for each language were then used to compare the base model against a fine-tuned version of the model as well as against the GPT-4 Turbo benchmark. In this case, the model was fine-tuned using a subset of the most challenging examples from the training dataset to enhance its performance on the task.

A second dataset of test examples was created by filling out the questionnaires by hand and then scanning them as PDFs using an iPhone 12 Mini smartphone camera. The scanned examples dataset was created so that it would be possible to explore how well the models generalized to different inputs. The dataset included 24 examples, i.e. three per language, and each scanned example was labeled manually to establish a ground truth structured representation. The manual labeling process was performed twice to ensure the veracity of the ground truth labels.

##### Figure S2: Structured Representation of a Questionnaire

Below is a JSON representation of a blank social needs questionnaire. It is worth noting that the template also included an instruction to the LLM to set the "language" correctly.

```
{
  "title": "CHA Social Needs Questionnaire",
  "language": "this must be the original language in which the input was written (must be in lowercase)",
  "questions": [
    {
      "text": "In the last 12 months, did you worry your food would run out before you got money to buy more?",
      "choices": [
        {
          "id": 1,
          "label": "Never",
          "state": "unselected"
        },
        {
          "id": 2,
          "label": "Sometimes",
          "state": "unselected"
        },
        {
          "id": 3,
          "label": "Often",
          "state": "unselected"
        }
```

```
        ]
    },
    {
        "text": "In the last 12 months, the food we bought didn't last and we didn't have money to get more.",
        "choices": [
            {
                "id": 4,
                "label": "Never",
                "state": "unselected"
            },
            {
                "id": 5,
                "label": "Sometimes",
                "state": "unselected"
            },
            {
                "id": 6,
                "label": "Often",
                "state": "unselected"
            }
        ]
    },
    {
        "text": "In the last 12 months, has the electric, gas or oil company threatened to shut off services in your home?",
        "choices": [
            {
                "id": 7,
                "label": "No",
                "state": "unselected"
            },
            {
                "id": 8,
                "label": "Yes",
                "state": "unselected"
            },
            {
                "id": 9,
                "label": "Already shut off",
                "state": "unselected"
            }
        ]
    },
    {
        "text": "In the last 12 months, have you or your family had trouble getting transportation to medical appointments?",
        "choices": [
            {
                "id": 10,
                "label": "No",
                "state": "unselected"
            },
            {
                "id": 11,
                "label": "Yes",
                "state": "unselected"
            }
        ]
    },
    {
        "text": "What is your living situation today?",
        "choices": [
            {
                "id": 12,
                "label": "I have a steady place to live.",
                "state": "unselected"
            },
            {
                "id": 13,
                "label": "I have a place to live today, but I am at risk of losing my housing.",
                "state": "unselected"
```

```
        },
        {
          "id": 14,
          "label": "I am temporarily staying with others, in a hotel, or a shelter.",
          "state": "unselected"
        },
        {
          "id": 15,
          "label": "I am living outside, in a car, abandoned building, or bus/train station.",
          "state": "unselected"
        }
      ]
    },
    {
      "text": "Do you have access to the internet at home? (Select all that apply)",
      "choices": [
        {
          "id": 16,
          "label": "Yes, on a computer",
          "state": "unselected"
        },
        {
          "id": 17,
          "label": "Yes, on my phone",
          "state": "unselected"
        },
        {
          "id": 18,
          "label": "No",
          "state": "unselected"
        }
      ]
    },
    {
      "text": "In the last 12 months, have you experienced violence at home or in your relationships? [Ambulatory Only]",
      "choices": [
        {
          "id": 19,
          "label": "No",
          "state": "unselected"
        },
        {
          "id": 20,
          "label": "Yes",
          "state": "unselected"
        },
        {
          "id": 21,
          "label": "Prefer not to say",
          "state": "unselected"
        }
      ]
    },
    {
      "text": "Would you like a CHA care team member to reach out to help you with the needs you checked off above?",
      "choices": [
        {
          "id": 22,
          "label": "Yes",
          "state": "unselected"
        },
        {
          "id": 23,
          "label": "No",
          "state": "unselected"
        }
      ]
    },
    {
```

"text": "Can we refer you to free or low-cost community programs (like food pantries) by sharing your name, phone, and address so they can reach you?",
"choices": [
{
"id": 24,
"label": "Yes",
"state": "unselected"
},
{
"id": 25,
"label": "No",
"state": "unselected"
}
]
},
{
"text": "Declined the questionnaire?",
"choices": [
{
"id": 26,
"label": "I do not want to answer these questions.",
"state": "unselected"
}
]
}
]
}

#### Analyzing the Text and Markdown Extracted from the Synthetic and Scanned Examples

The questionnaire contains 26 checkboxes, so it would be expected that the text and markdown representations of the questionnaire generated by the Document Intelligence Layout API would contain the same number of selection mark tokens. In the synthetic dataset, the API detected exactly 26 selection mark tokens while detecting 27 tokens in the examples in other languages. The extra selection mark token was likely due to the check mark in the questionnaire's instructions confusing the model. In the scanned dataset, the number of selection mark tokens in the text and markdown representations were also somewhat inconsistent ranging from 26 to 28 tokens. Lastly, it is important to note the number of selection mark tokens was consistent between the text and markdown representations.

For more detail, please see:

- **Analyzing Synthetic Dataset:** https://oekekezie.github.io/poc-ai-sdoh-questionnaire-v2/html/Analyzing%20Synthetic%20Dataset.html
- **Analyzing Scanned Dataset:** https://oekekezie.github.io/poc-ai-sdoh-questionnaire-v2/html/Analyzing%20Scanned%20Dataset.html

#### Fine-Tuning GPT-3.5 Turbo

The fine-tuned GPT-3.5 Turbo model with 128 examples is referred to as “small” while the one that was trained on 256 examples is referred to as “large.” All of the fine-tuning jobs used the same seed. The fine-tuning hyperparameters included batch size (the number of training examples processed before the model's internal parameters are updated), learning rate (the size of the update to the model's weights each time they are adjusted), and the number of epochs (the total number of times the complete dataset is passed through the model during training). The optimal fine-tuned models were selected (highlighted in yellow) based on demonstrating the lowest full validation loss (computed on the entire validation dataset at the end of each epoch, providing a comprehensive view of the model's overall performance) and for which the validation loss (calculated on small batches of validation data during each training step, offering iterative insights into the model's performance) most stably decreased over the epochs:

##### Table S1: Fine-Tuning Job Hyperparameters

| OpenAI Model | Training (N) | Validation (N) | Trained Tokens | Epochs | Batch Size | Learning Rate Multiplier | Training Loss | Full Validation Loss |
|---|---|---|---|---|---|---|---|---|
| gpt-3.5-turbo-0125 | 104 | 24 | 1,001,469 | 3 | 1 | 2 | 0 | 0.0306 |
| gpt-3.5-turbo-0125 | 104 | 24 | 1,001,469 | 3 | 1 | 0.5 | 0 | 0.0455 |
| gpt-3.5-turbo-0125 | 104 | 24 | 1,001,469 | 3 | 1 | 0.1 | 0.1011 | 0.0602 |
| gpt-3.5-turbo-0125 | 104 | 24 | 1,001,469 | 3 | 4 | 2 | 0.0032 | 0.0022 |
| gpt-3.5-turbo-0125 | 104 | 24 | 1,001,469 | 3 | 8 | 2 | 0.0031 | 0.0026 |
| gpt-3.5-turbo-0125 | 104 | 24 | 1,669,115 | 5 | 8 | 0.5 | 0.0026 | 0.0038 |
| **gpt-3.5-turbo-0125 (Small)** | **104** | **24** | **1,669,115** | **5** | **8** | **2** | **0.0005** | **0.0016** |
| **gpt-3.5-turbo-0125 (Large)** | **208** | **48** | **3,336,280** | **5** | **8** | **2** | **0.0004** | **0.0015** |
| gpt-3.5-turbo-0125 | 208 | 48 | 6,672,560 | 10 | 8 | 2 | 0 | 0.0016 |

#### Comparing Performance on Synthetic Testing Dataset

Please see:
https://oekekezie.github.io/poc-ai-sdoh-questionnaire-v2/html/Comparing%20Performance%20on%20Test%20Dataset.html

#### Comparing Performance on Scanned Testing Dataset

Please see:
https://oekekezie.github.io/poc-ai-sdoh-questionnaire-v2/html/Comparing%20Performance%20on%20Scanned%20Dataset.html
